# A web-based Diagnostic Tool for COVID-19 Using Machine Learning on Chest Radiographs (CXR)

**DOI:** 10.1101/2020.04.21.20063263

**Authors:** E.B. Gueguim Kana, M.G. Zebaze Kana, A.F. Donfack Kana, R.H Azanfack Kenfack

## Abstract

This paper reports the development and web deployment of an inference model for Coronavirus COVID-19 using machine vision on chest radiographs (CXR). The transfer learning from the Residual Network (RESNET-50) was leveraged for model development on CXR images from healthy individuals, bacterial and viral pneumonia, and COVID-19 positives patients. The performance metrics showed an accuracy of 99%, a recall valued of 99.8%, a precision of 99% and an F1 score of 99.8% for COVID-19 inference. The model was further successfully validated on CXR images from an independent repository. The implemented model was deployed with a web graphical user interface for inference (https://medics-inference.onrender.com) for the medical research community; an associated cron job is scheduled to continue the learning process when novel and validated information becomes available.

## Introduction

The novel coronavirus outbreak was first identified in December 2019 in Wuha, in Hubei province, China. It is caused by Severe Acute Respiratory Syndrome Coronavirus 2 (SARS-CoV-2)(Stoecklin *et al*., 2020). The virus has a zoonotic origin and, earlier in 2007 Chen *at al*., pointed out that the presence of a large reservoir of SARS-CoV-like viruses in horseshoe bats, together with the culture of eating exotic mammals in southern China, was a time bomb.

The virus is mainly spread through close contact and via respiratory droplets (cough or sneeze) and through fomites (Centers for Disease Control and Prevention (CDC ^a^), 2020). Patients with COVID-19 show clinical manifestations including fever, non-productive cough, dyspnoea, myalgia, fatigue, normal or decreased leukocyte counts, and radiographic evidence of pneumonia (Huang, 2019). The Virus has spike-like S proteins surrounding its envelope structure which mediates its attachment to specific receptors on lung cells called the ACE2 receptors using a “key and lo ck” tactic, facilitating viral entry into the host cell (Tai, 2019). This leads to an inflammatory process in the lungs, causing shortness of breath and in more severe cases, there is a release of cytokines in the bloodstream, thus damaging vital organs.

An exponential spread of this disease has since been observed globally, and the World Health Organization (WHO) declared it as a Public Health Emergency of International Concern on 30 January 2020, and subsequently recognized COVID-19 as a pandemic on 11 March 2020 (WHO, 2020). In the absence of vaccine, and proven drugs to control the pandemic and prevent the collapse of global care system, WHO has recommended countries to isolate, test, treat and trace.

Radiology observations of COVID-19 infected individuals show bilateral multifocal consolidations (fluid or other products of inflammation filling pulmonary air space) that may progress to involve entire lungs. There is also a presence of small pleural effusions which is an abnormal fluid developed in the spaces around the lungs (Kampalath, 2020). Thus there is presence of glass patterned areas, which affect both lungs even in the initial stages of the infection, in particular the lower lobes, and especially the posterior segments, with a fundamentally peripheral and subpleural distribution (Figure 1).

**Figure 1:**
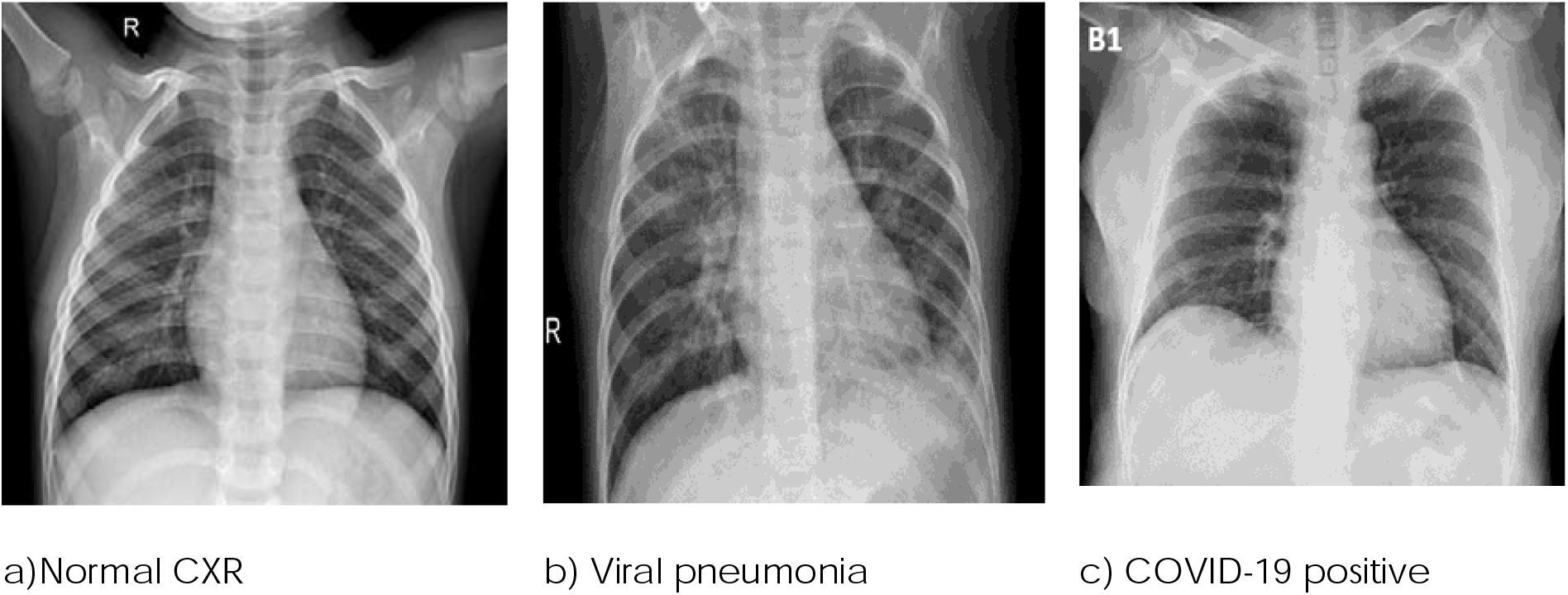
CXR images of (a) healthy individual, (b) other Viral or Bacterial pneumonia, (c) COVID-19 positive patient

Currently, to detect and diagnose an infected individual upon clinical suspicion, or monitor the disease, a real-time reverse transcription polymerase chain reaction (rRT-PCR) test is done (CDC ^b,^ 2020) followed by chest radiographs (CXR) and/or computed tomography (CT) scans. rRT-PCR test results may take several hours or days and there is a pronounced shortage of RT-PCR tests kit, unlike the chest CXR which are routinely available with an instant outcome. Italian and British hospitals are beginning to employ CXR as a first-line triage tool due to long reverse transcription polymerase chain reaction (RT-PCR) turnaround times (Wong et al., 2020). There is a growing interest in the use of CXR and CT scans for the screening, diagnosis and management of patients with suspected or known COVID-19 infection. CT scans are more sensitive than RT-PCR, thus a potential advantage of using them (Milagros, 2020). However, the interpretation of CXR and CT-scan is laborious, time-consuming and requires radiologist expertise. An immediate access to such expertise may not always be available in all clinical settings. These challenges could be addressed with computer vision.

### Artificial intelligence image vision in medical diagnosis

There is an increasing interest in the use of Artificial Intelligence (AI) technologies to support disease diagnosis and management in areas of ophthalmology, pathology, cancer detection, radiology or prediction and personalized medicine. The use of Convolutional Deep Neural Network (CDNN) allows for impressive discovery of patterns in medical image datasets, achieving image classification sometimes difficult for human experts (Zhou et al., 2019). The CDNN uses multiple processing layers to learn and represent data with multiple levels of abstraction by applying image analysis filters, or convolutions. The abstracted representation of images within each layer is constructed by convolving multiple filters across the image, producing a feature map that is used as input to the next layer. Thus with this architecture, the image pixels are used as input, then processed to generate the desired classification output. The development and use of Graphical Processing Unit (GPU) instead of CPU and availability of large open-source image datasets for model development have fuelled advances in machine vision (Athanasio et al., 2018).

This study aimed at developing and deploying an inference model for rapid detection of COVID-19 using transfer learning and Machine vision on chest radiographs (CXR).

## Materials and Methods

### Data source and transformation

A total of 2507 bacterial or viral pneumonia infected, 2487 healthy individuals and 161 COVID-19 positives CXR images were sourced from two repositories and used for model development (Kermany *et al*., 2018; Cohen, 2020). Additional 2585 background class images labelled as “Not Certain” were included. To address the challenge of class imbalance for COVID-19 positive (1:13), data augmentation through contrast transformation (scale: 0.5-1.9) with probability of 0.5 was invoked for this class to achieve a total of 2093. Further data augmentation such as random rotation, horizontal flipping, horizontal and vertical shifting and magnification were not considered for this dataset. Thus a total of 4 classes of healthy individual, bacterial and Viral pneumonia, COVID-19 infected and “Not certain” with 7254 training and 2418 validations image were used. These images were reviewed for their quality, then sized to 350 pixels and normalized according to imagenet statistics.

### Model development

The transfer learning of Resnet50 architecture was leveraged for model development. Resnet50 has shown a high performance on benchmark dataset such as Imagenet dataset, a huge database of labelled images containing more than 14 million images which have been hand-annotated and distributed into more than 20,000 classes suitable for training Convolutional Neural Networks. Transfer learning has proven to be a highly effective approach which addresses the limitation in data sizes and lowers the computing cost (Yosinski et al., 2014). It leverages previous knowledge acquired while solving one problem and applying it to a different but related problem (Wang et al., 2020). Thus, through adopting Restnet architecture, the optimized weights gained from previous, and more general trainings were fixed in the lower layers, and only the weights of the upper layers were retrained on chest radiographs (Figure 2).

The model was developed using python language and trained on the subset of training data with differential learning rate (10^−4^ - 10^−2^) for 15 epochs on Tesla K80 GPU from google cloud resources. Each epoch corresponded to one complete presentation of the training set to the model.

**Figure 2:**
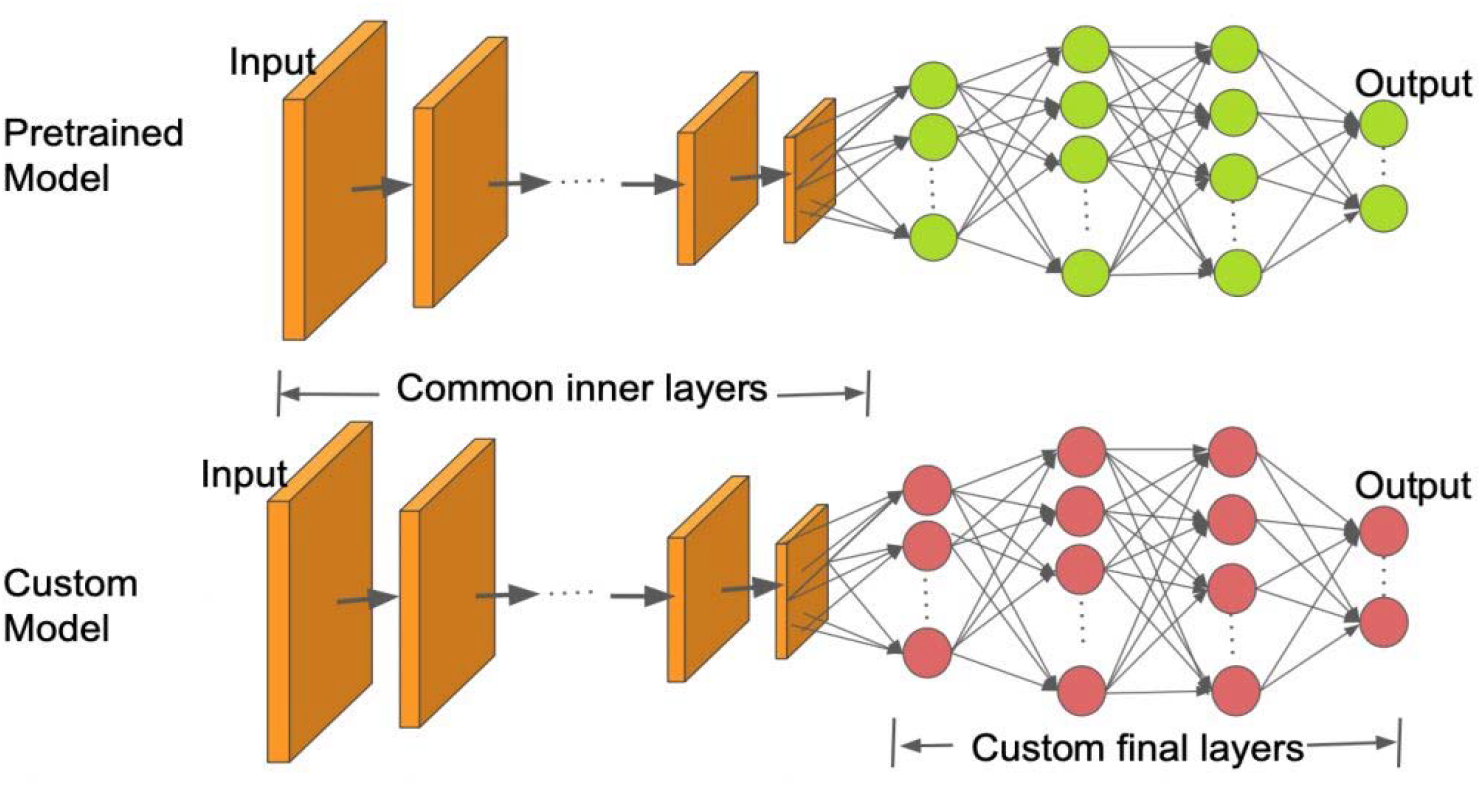
Illustrative diagram of transfer learning using pre-trained model (Nayak, 2019).

### Model performance metrics

The performance of the model was assessed using the accuracy, precision, recall and F-score. The precision metric is particularly useful when the false positives are of great concern. The recall is important when the impact of false negatives on decision making is high. These were computed using Equations 1 and 2 respectively (Olson, 2008). In some instances, the observed accuracy could be largely contributed by the vast number of true negatives, which may be acceptable for some classes of business decision, whereas false negative and false positive could pose irreversible damage in medical decision, thus F-score was computed using Equation 3.

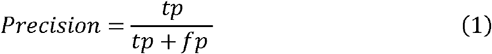

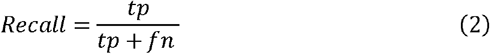

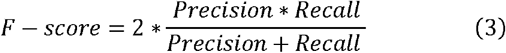

### Model validation on external independent dataset and deployment

The developed model was further validated on a sample data from an additional repository of CXR images (Chowdhury et al., 2020). This repository was curated by expert researchers and medical practitioners consisting of 219 COVID-19 positive images, 1341 normal images and 1345 viral pneumonia images. From this dataset, random samples of 200 images per class were validated on the developed model. The developed model was deployed on the cloud with a web GUI for CXR inference.

## Result and Discussion

### Model development and performance metrics

The availability of a large labelled dataset of medical images required for the machine vision model development remains a critical challenge due to privacy concerns in data sharing, and the laborious task of data labelling. Transfer learning enables model development using fewer datasets. In this study, Resnet was trained on fewer instances (9672 images) for 30 epochs to obtain an accuracy of 99%. Training was stopped to avoid overfitting as there were no further indications of learning.

The confusion matrix is depicted in Figure 4 based on unseen images for a bacterial or viral pneumonia, out of 2418 instances of images, 2383 were accurately predicted (true positive) and only 35 were wrongly predicted (false positives and false negatives). For COVID-19 class, out of 540 validation instances, 539 were accurately predicted (true positive) and 1 was wrongly predicted as viral or bacterial pneumonia. Zu et al., (2019) reported that in patients with severe disease, their X-ray readings may resemble pneumonia or acute respiratory distress syndrome (ARDS). Notwithstanding, in the case of COVID-19, a single case of a false-negative outcome for COVID-19 infected individual is a serious concern as it could compromise global efforts to contain the pandemic; similarly, a false positive can cause undue psychological distress, possible contamination if quarantined with infected individuals or unnecessary investigation which places extra burdens on the healthcare system. However, with the current COVID-19 diagnostic procedure, radiography results are used to support the clinical symptomatic examination and could be further confirmed through PCR results. The associated precision and recall values for each class are depicted in table 1.

**Table 1:** Performance of the classifier

| Class | Precision | Recall | Accuracy | F1 measure |
| --- | --- | --- | --- | --- |
| bacterial or viral pneumonia | 98.8% | 97.2% | 98.8% | **98%** |
| COVID-19 Positive | 100% | 99.8% | 100% | **99.8%** |
| healthy individual | 95.6% | 99% | 95.6% | **97.3%** |
| Not Certain | 100% | 100% | 100% | **100%** |
| Average | **99%** | **99%** | **99%** | **99%** |

**Figure 3:**
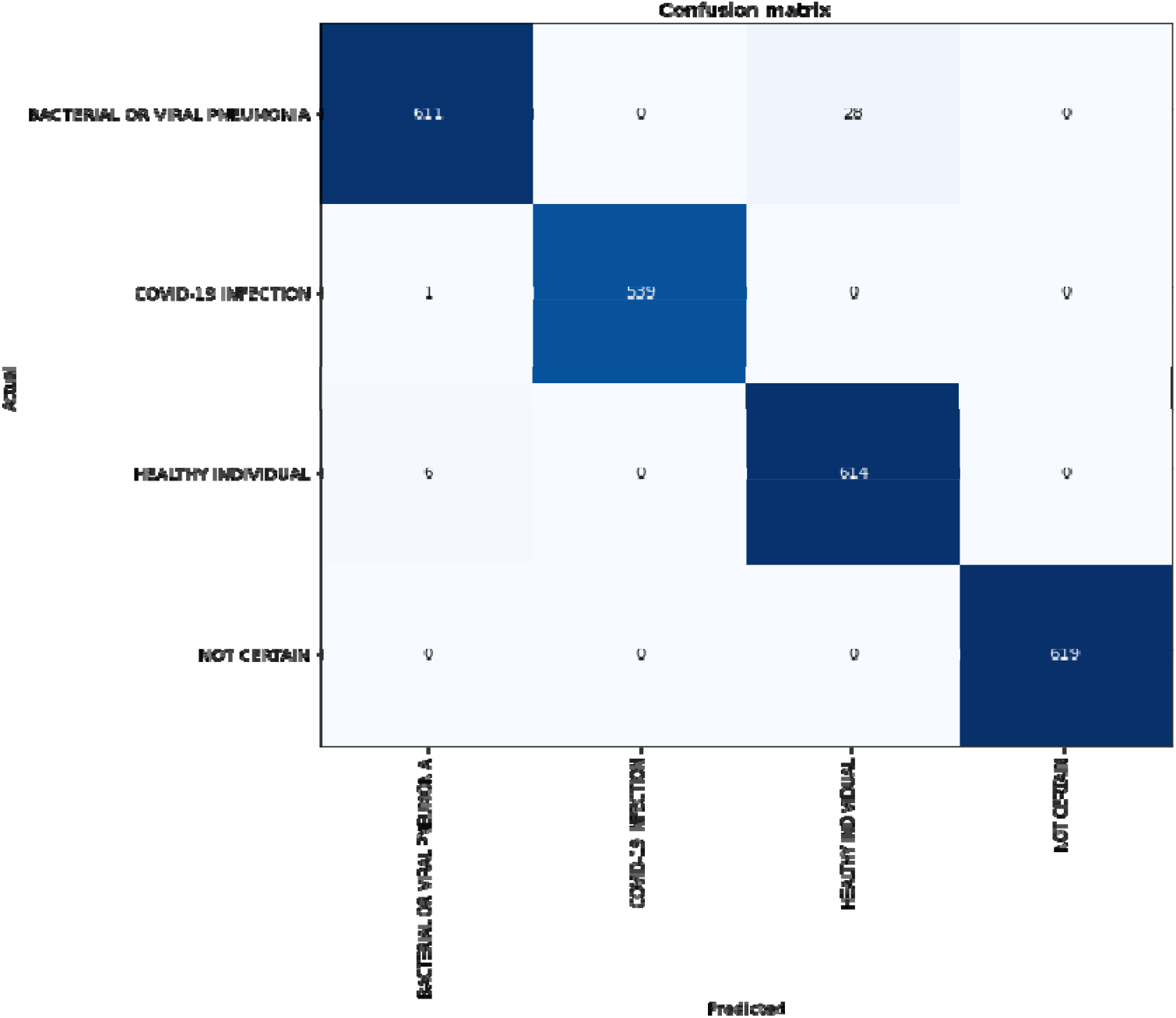
Confusion matrix from validating subset of data.

**Fig 4.**
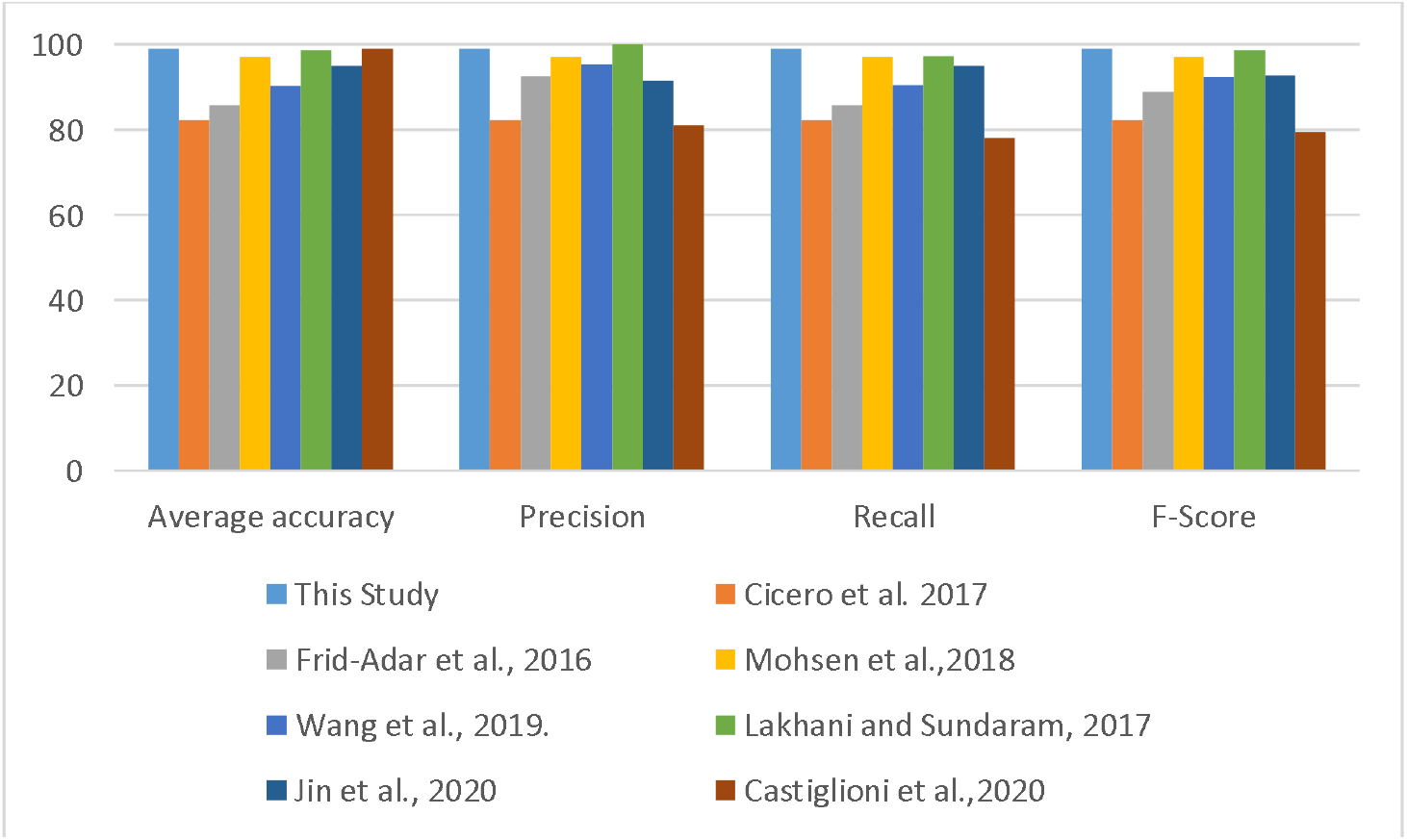
Comparison chart for the performance of classifiers.

A false-positive result occurs when an individual is inaccurately assigned to a class, such as a healthy individual categorized as COVID-19 patient and a false negative occurs when an individual who accurately belongs to a given class is excluded from such group.

### Model performance on an additional independent dataset

When the developed model was assessed on CXR data from an additional repository using a subset of 600 CXR images of COVID-19 positives, viral or bacterial pneumonia and normal persons (200 images per class), accuracies of 97%, 85% and 98% respectively were observed.

### Comparison with some existing models

A comparison of the developed model with other reported machine vision models for medical image inference is shown in Table 2 and the corresponding chart for the performance of classifiers is displayed in Figure 4. A noticeable trend in the use of machine vision for medical diagnosis has been observed in the last few years as evidenced in Table 2. The purpose of this comparison was to highlight the similarities in performance metrics and effectiveness of the developed COVID-19 model with other reported machine vision models for medical image inference.

**Table 2.**
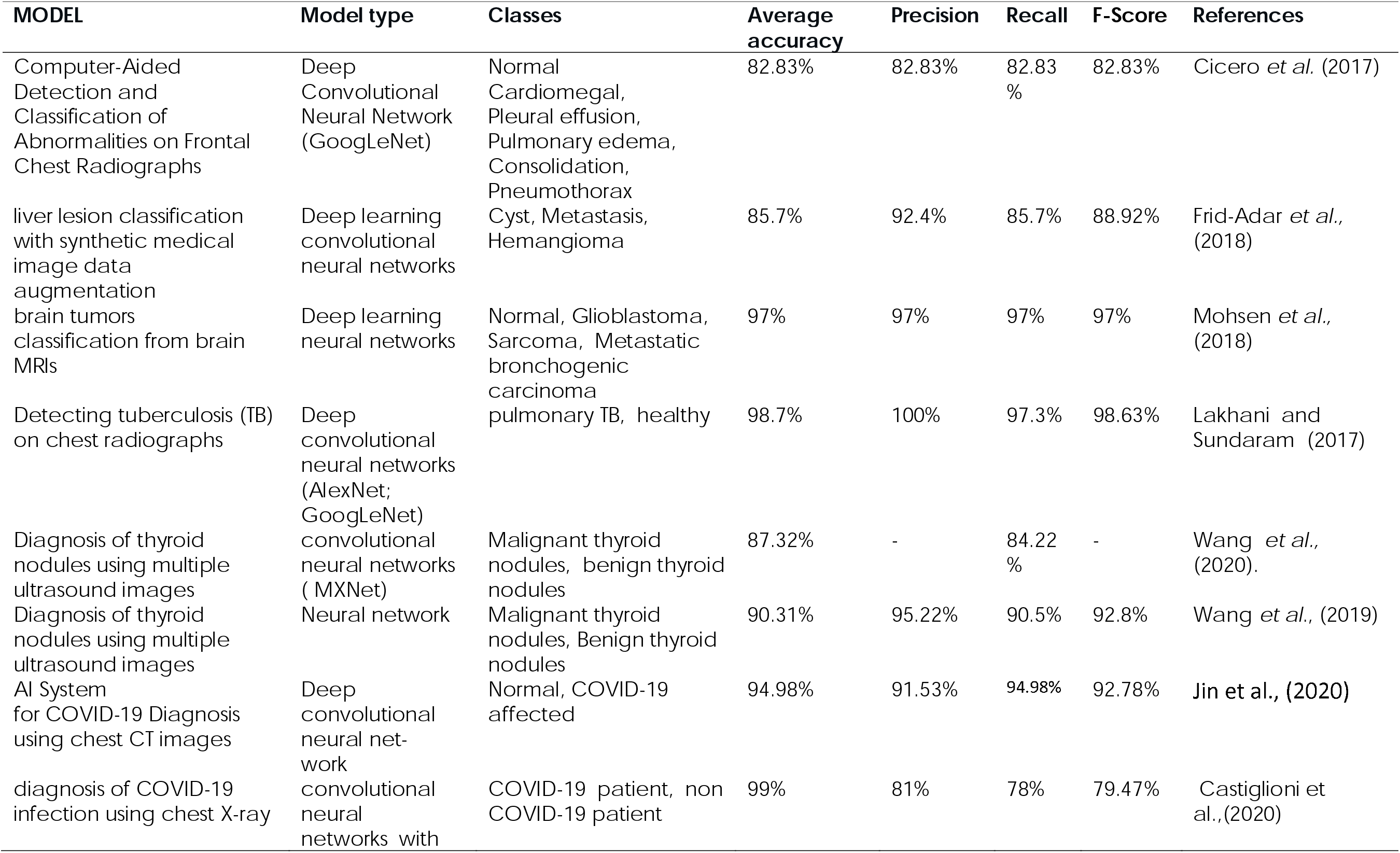
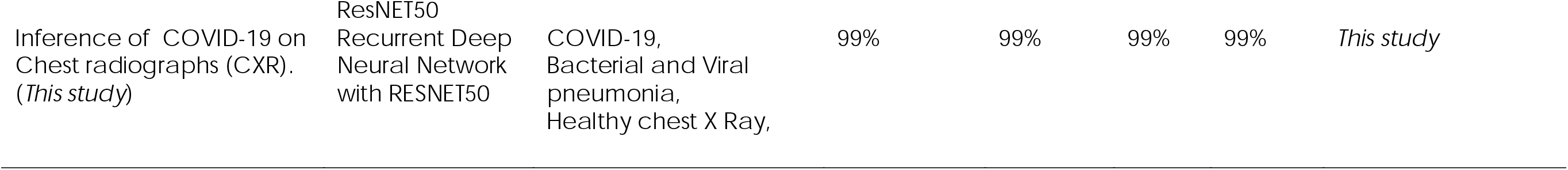
Model comparison with some reported AI models on medical images.

The observed a ccuracy rate, recall, precision, and F-Measure for all the four classes (COVID-19, bacterial and Viral pneumonia, Healthy chest X Ray, Not Certain) varied from, 95-100%, 98-100%, 97-100% and 97-100% respectively, and average values of 99% were observed for all the metrics. These performance values compared effectively or slightly better than other reported classifiers in the same terms (Table 2 and Figure 4). Using Deep neural network to diagnose pulmonary tuberculosis, Lakhani and Sundaram (2019) reported performance metrics values of 98.7%, 100%, 97.3% and 98.63% for accuracy, precision, recall and F-score respectively. In the same vein Cicero et al. (2017) investigated the suitability of Deep Convolutional Neural network for detection and classification of abnormalities on frontal chest Radiographs and achieved performance values of 82.83%, 82.83%, 82.83%, 82.83% for accuracy, precision, recall and F-score respectively. In the diagnosis of COVID-19, Jin et al., (2020) used Deep convolutional neural network on chest CT images and obtained 94.98%, 91.53%, 94.98%, 92.78% for accuracy, precision, recall and F-score respectively. Similarly, Castiglioni et al.,(2020) used convolutional neural networks on chest X-ray and obtained 99%, 81%, 78%, 79.47% for accuracy, precision, recall and F-score respectively. These observations highlight the potential of the developed COVID-19 model to accurately discern the disease pattern on novel CXR images.

### Significance of the study for COVID-19 diagnosis

The model’s ability to recognise pixels of glass patterned areas on CXR as distinguishing features of COVID-19 and to differentiate these from other viral and pneumonia infected lungs or healthy images resulted from the knowledge gained from the repeated exposures of the neural network to positive training instances, thus providing prediction similar to human experts.

With the rapidly expanding COVID-19 pandemic, the demand for chest radiographs has grown exponentially and proportionally with the number of patients visiting the emergency departments (Milagros, 2020) thus generating a large number of CRX images. Immediate access to expert interpretation may not available or affordable in all clinical settings, thus computer-assisted decision support may be useful to clinicians and radiologists, and for prioritizing cases in a large worklist.

Furthermore, considering that if a massive screening of the population is performed as recommended by the WHO in attempts to curb the infection curve, assuming only 5% the global population (7.8 billion inhabitants) is eventually screened (Worldometer,2020) that would generate 390 million images which need to be reviewed. This will place an unprecedented burden on human experts. Thus automated image analysis tool based on machine learning algorithm, with friendly GUI (Figure 5) will be a key enabler to meet this goal

**Figure 5:**
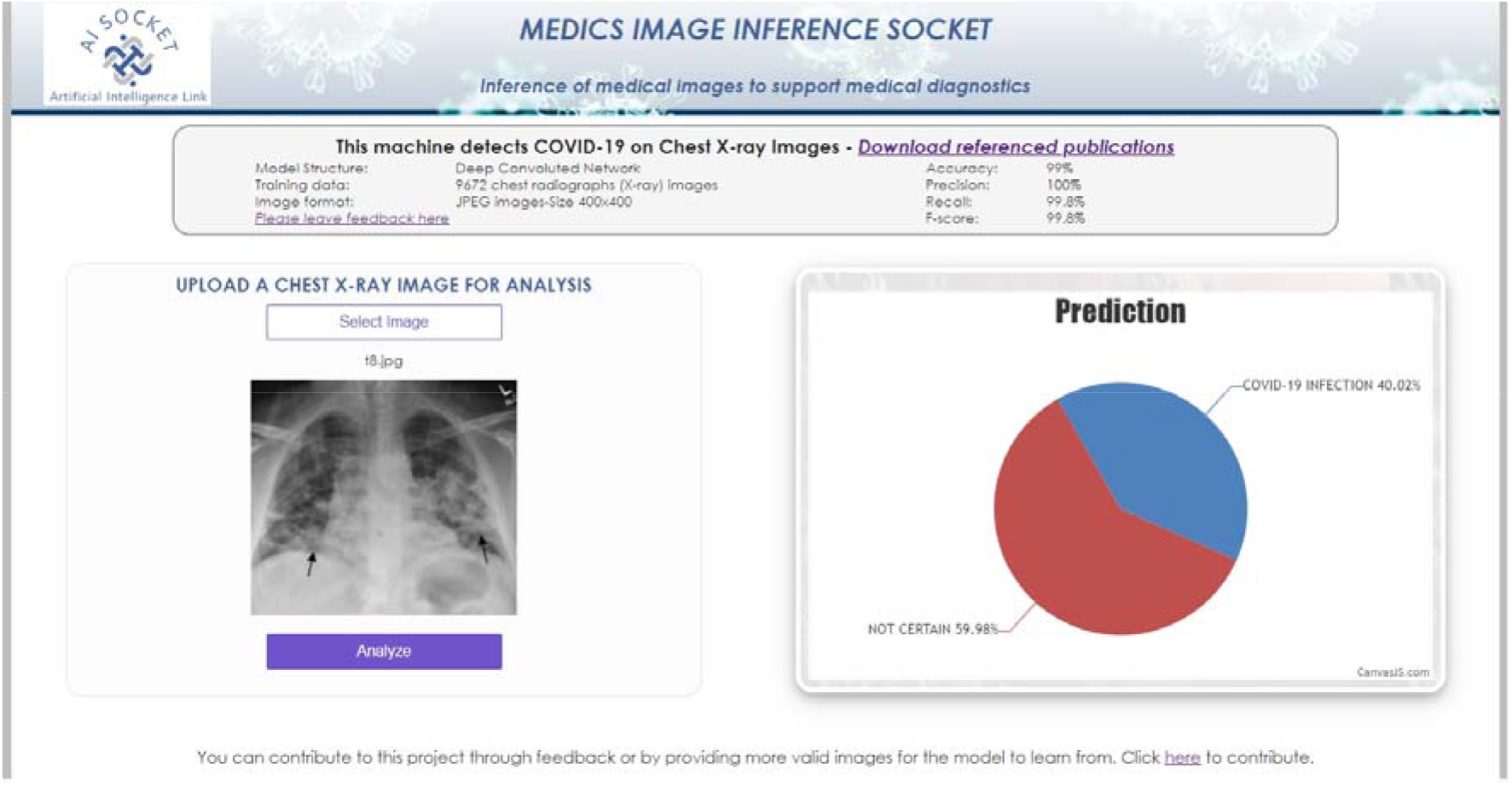
Web graphical User Interface (GUI)

## Data Availability

The links for the data used in the model development have been provided in the manuscript

